# Symptomatic hypermobility as a risk factor for Long COVID with high post-exertional symptom exacerbation: further analysis of data from a retrospective online survey of adults in the United States and United Kingdom

**DOI:** 10.64898/2026.06.24.26356475

**Authors:** Jeffrey Lubell, Regina A. Torok, Rena M. Rudy, Lisa Quadt, Jessica A. Eccles

**Author notes:** These authors contributed equally to this manuscript as first authors (Jeffrey Lubell, Regina A. Torok). These authors contributed equally to this manuscript as senior authors (Lisa Quadt, Jessica A. Eccles). **Correspondence to: Dr Jessica Eccles**, Department of Neuroscience, Brighton and Sussex Medical School, Trafford Centre, University of Sussex, BRIGHTON BN1 9RY.

## Abstract

**Background:** In a retrospective online survey, we assessed the extent to which people with symptomatic hypermobility are at risk of Long COVID with a high degree of post-exertional symptom exacerbation, a form of Long COVID similar to myalgic encephalomyelitis.

**Methods:** Participants were 1,816 adults with prior COVID-19 infection; 19.4% reported Long COVID, defined as symptoms persisting ≥3 months. Survey measures identified Long COVID with high post-exertional symptom exacerbation, generalized joint hypermobility (GJH), extreme hypermobility, and a pre-COVID orthostatic/neurocognitive symptom burden (ONS profile). Logistic regression assessed whether ONS profile and hypermobility, together defined as symptomatic hypermobility, were associated with increased risk of Long COVID with post-exertional symptom exacerbation.

**Results:** In the full sample, both extreme hypermobility (OR 3.15, 95 % CI 2.00-4.95) and an ONS profile pre-COVID (OR 3.29, 95% CI 2.34-4.61) were strongly predictive of Long COVID with high post-exertional symptom exacerbation. These effects were cumulative, leading to an OR of 9.46 (95% CI 4.93-18.17) for people with both conditions. People who both had an ONS profile pre-COVID and had generalized joint hypermobility also had a higher risk of Long COVID with high post-exertional symptom exacerbation (OR 5.54, 95% CI 3.51-8.75).

**Conclusions:** In this dataset, people with symptomatic hypermobility were at high risk of Long COVID with high levels of post-exertional symptom exacerbation. Further research is needed to understand the biological mechanisms of viral-onset illness to promote more effective and targeted treatments tailored to the disease pathways shared by groups of individuals with common vulnerabilities.

**Summary:** This paper uses data from a retrospective online survey to assess the extent to which people with symptomatic hypermobility are at risk of Long COVID with post-exertional symptom exacerbation, a form of Long COVID similar to myalgic encephalomyelitis.

---

In an earlier analysis of data from an online survey of 1,816 adults in the United States and United Kingdom (352 reporting Long COVID and 1464 reporting no Long COVID), we showed that both generalized joint hypermobility (GJH) and extreme hypermobility were significant risk factors for developing Long COVID.^1^ Other research has similarly found an association between GJH and Long COVID.^2,3^ In this paper, we refine our earlier analysis by assessing the extent to which people with symptomatic GJH and symptomatic extreme hypermobility are at risk of a Long COVID presentation that approximates myalgic encephalomyelitis (ME; sometimes referred to as myalgic encephalomyelitis/chronic fatigue syndrome; ME/CFS): Long COVID with a high degree of post-exertional symptom exacerbation (PESE).

People with GJH have joints that move beyond the normal range, often due to lax ligaments from variant connective tissue in the body. GJH is estimated to affect approximately 10 to 20% of the general population,^4^ with rates up to 78.8% in people with complex chronic conditions, including ME.^5^

Not everyone with GJH develops symptoms related to their hypermobility. S*ymptomatic GJH*^*c*^ is characterised by musculoskeletal manifestations such as chronic or widespread pain,^7–9^ recurrent subluxations, joint instability, and musculoskeletal overuse injuries (including tendinitis, bursitis).^10^ Approximately two-thirds^6^ of people with *symptomatic* GJH^11^ also report multisystemic symptoms,^12^ including physical and cognitive fatigue,^7,8,13,14^orthostatic intolerance^15,16^ and/or postural orthostatic tachycardia syndrome (POTS),^16,17^ along with gastrointestinal symptoms^18^ and features consistent with mast cell activation syndrome.^19^ Many people with symptomatic hypermobility are diagnosed with hypermobile Ehlers-Danlos Syndrome (hEDS) or another less common form of Ehlers-Danlos Syndrome, or one of several hypermobility spectrum disorders (HSD). The estimated combined diagnosed prevalence of hEDS/HSD is between 1 in 500 to 1 in 900 individuals,^20,21^ though some argue it is inconsistently diagnosed.^11^

A number of studies have found high rates of hEDS, HSD and GJH among people with ME,^22–25^ suggesting that people with symptomatic hypermobility may be at higher risk of developing this severe chronic condition. Although the extent to which ME and Long COVID overlap remains debated, growing evidence suggests that many people with long COVID have a condition closely resembling, or effectively identical to, ME.^1,26,27^ One of the signature characteristics of ME is post-exertional neuroimmune exhaustion,^28^ often referred to as post-exertional malaise. While our dataset is not large or comprehensive enough to assess whether symptomatic hypermobility is a risk factor for the multifaceted post-exertional neuroimmune exhaustion, we are able to assess whether people with hypermobility who were burdened before getting COVID by some of the systemic symptoms often experienced by people with hEDS are at greater risk of Long COVID with PESE. PESE is a worsening of symptoms after exertion and is one of the constituent components of post-exertional neuroimmune exhaustion.

In this paper, we examine whether having a high burden from orthostatic or neurocognitive symptoms pre-COVID plus either GJH or extreme hypermobility predicts Long COVID and Long COVID with high PESE. In addition to considering the results for the full sample, we examine the results for two sub-groups defined by the severity of initial symptoms during the acute phase of COVID-19. This analysis contributes to the field’s understanding of the clinical relevance of symptomatic hypermobility, while also raising important questions about the pathophysiology of ME-type presentations of Long COVID.

## Methods

### Setting

This analysis is based on data collected through a survey conducted online via the platform Qualtrics in early 2024. The study was approved by the Research Ethics and Governance Committee of the Brighton and Sussex Medical School (ER/BSMS9B02/6/4).

### Public and Patient Involvement

The authorship team includes individuals with hypermobility and Long COVID, as well as a patient caregiver. The questionnaire was refined using the suggestions of additional individuals with Long COVID.

### Participants

The dataset includes survey responses from 1,816 adults in the United States and United Kingdom who reported at least one COVID-19 infection at least three months before completing the survey. 352 respondents reported Long COVID and 1464 reported no Long COVID. We recruited respondents from representative online panels maintained by the data firm Dynata. For more details on the survey and survey recruitment, see Torok et al. 2025.^1^

### Outcomes

#### Long COVID

We identified respondents as having Long COVID based on their self-identification in response to the survey question “Have you been diagnosed with Long COVID, or do you have symptoms that you attribute to Long COVID? For the purposes of this survey, people have Long COVID when they experience symptoms related to their COVID-19 infection at least three months after their initial infection.”

#### Long COVID with high PESE

We identified Long COVID with high PESE based on responses to the following question asked of people who self-identified as having Long COVID: “Do you CURRENTLY experience a worsening of your symptoms up to 48 hours after you engage in even a small amount of exertion (e.g., doing basic housework)?” Responses were indicated by a slider from 0 (“not a problem) to 100 (“an extremely severe problem.”) We categorized respondents who had Long COVID and provided a response to this question at or above the median response (for those with Long COVID) of 20.5 as having Long COVID with high PESE.

### Subgroups

#### GJH and Extreme Hypermobility

We identified respondents as having GJH using the Hakim-Grahame self-report questionnaire (5PQ), where a score of 2 or greater indicates presence of GJH. We assessed extreme hypermobility by summing the number of affirmative responses to 9 questions – the 5PQ plus 4 additional questions. We identified respondents in the top 10^th^ percentile for their age and gender category as having extreme hypermobility. See Torok et. al. 2025 for more details.^29^ All of the individuals with extreme hypermobility also have GJH.

#### ONS Profile Pre-COVID and Symptomatic Hypermobility

We assessed the extent to which respondents report being burdened before getting COVID-19 by one of the orthostatic or neurocognitive symptoms commonly experienced by people with hEDS (ONS profile). We identify respondents as having an ONS profile pre-COVID if their self-reported pre-COVID-19 levels of any of four specific symptoms – fatigue, dizziness, cognitive impairment or inability to remain upright – were in the top tenth percentile of survey responses. These questions used a slider and were phrased similarly to the question about PESE referenced above. We identified respondents as having symptomatic GJH if they had both an ONS profile pre-COVID and GJH. We identified respondents who had an ONS profile pre-COVID and had extreme hypermobility as having symptomatic extreme hypermobility. We use the phrase symptomatic hypermobility to refer to respondents with either symptomatic GJH or symptomatic extreme hypermobility.

Figure 1 shows the relationship between having an ONS profile Pre-COVID and having GJH or extreme hypermobility.

**Figure 1:**
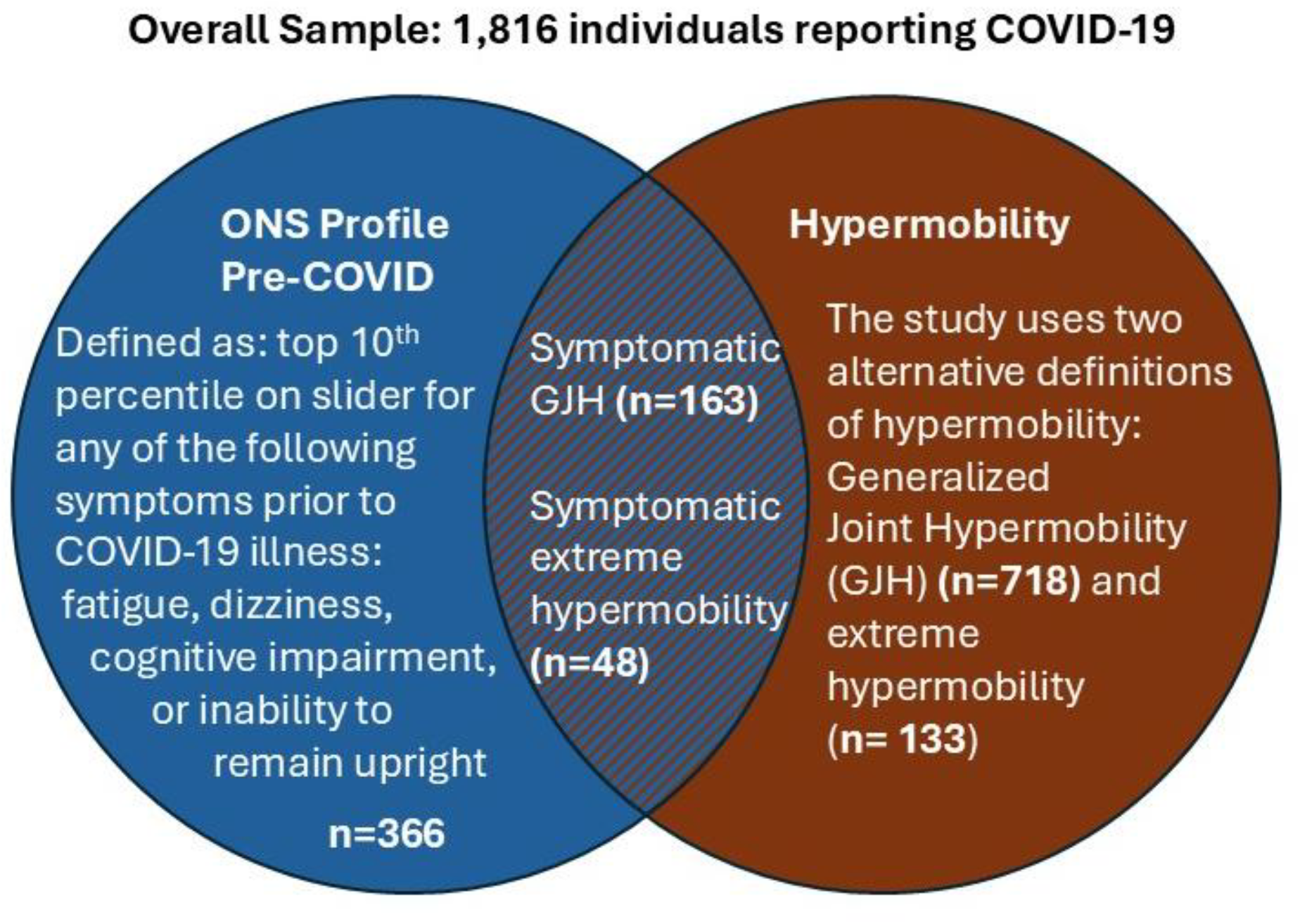
Venn Diagram of Symptom Categories.

#### COVID-19 Symptoms During Active Infection(s)

In addition to examining whether and to what extent respondents reported having an ONS profile *prior* to their COVID-19 infection, which we used to identify individuals with symptomatic GJH or symptomatic extreme hypermobility, we also used respondents’ reports of any symptoms *during* the acute phase of their COVID-19 infection(s) to create subgroups for analysis. One subgroup consists of individuals who reported no or mild symptoms during (all of) their COVID-19 infection(s); the other subgroup consists of individuals who reported severe symptoms during at least one COVID-19 infection.

### Statistical Analysis

We used Jamovi version 2.3.28 and IBM SPSS Statistics version 29 to analyze unweighted data.

### Descriptive Data

We tabulated descriptive data on the extent to which people in the full sample and three outcome categories – individuals reporting no Long COVID, individuals reporting Long COVID without high PESE and individuals reporting Long COVID with high PESE – report GJH, extreme hypermobility, an ONS profile pre-COVID, symptomatic GJH and symptomatic extreme hypermobility. We used chi-square tests to assess the significance of deviations from expected values (based on results for the full sample).

### Logistic Regressions

We used a series of separate binomial logistic regressions to test our hypothesis that hypermobility and symptomatic hypermobility are predictive of Long COVID with high PESE. Each of these regressions controlled for age, sex at birth, number of infections (up to 3), and number of vaccine doses, factors previously shown to affect the risk of Long COVID.^30^

We conducted four separate regressions of the full sample; each regression had Long COVID with high PESE as the dependent variable and included the common covariates noted above plus additional covariates noted below:

The first regression included these predictors: GJH and ONS profile pre-COVID. We also included the interaction term of these two predictors because it was significant (before Bonferroni correction).

The second regression included these covariates: symptomatic GJH, ONS profile pre-COVID but without GJH, and GJH without ONS profile pre-COVID. This allowed for an estimation of the impact of symptomatic GJH.

The third regression included these covariates: extreme hypermobility and ONS profile pre-COVID. We excluded the interaction term of these two covariates because it was not statistically significant.

The fourth regression included these covariates: symptomatic extreme hypermobility, ONS profile pre-COVID but without extreme hypermobility, and extreme hypermobility without ONS profile pre-COVID. This allowed for an estimation of the impact of symptomatic extreme hypermobility.

We repeated these four regressions for each of two subgroups: respondents reporting mild or no initial symptoms during (all of) their COVID-19 infection(s) and respondents reporting severe initial symptoms during (any of) their COVID-19 infection(s). This yielded a total of 12 separate binomial regressions with Long COVID with high PESE as the dependent variable. Because the interaction term in the first regression was significant for the full sample (before Bonferonni correction), we included it in the first regression for each of the subgroups as well.

We also conducted the same 12 binomial regressions using Long COVID as a dependent variable, rather than Long COVID with high PESE. We assessed the interaction terms but omitted them because they were non-significant in all regressions.

We corrected for multiple comparisons using a Bonferroni correction (p=0.05/12 equals the corrected significance of p=0.004).

We also converted the odds ratios associated with developing Long COVID with high PESE to risk ratios using standard formulas. We computed the estimated incidence of Long COVID with high PESE for people with symptomatic GJH and symptomatic extreme hypermobility by multiplying the risk ratios by the percentages for the comparison populations. These incidences thus reflect the regression-adjusted incidence of Long COVID with high PESE, rather than the unadjusted observed incidence for these subgroups. The comparison groups for the risk ratio calculations are people without an ONS profile pre-COVID and without GJH (in one set of risk ratios) or extreme hypermobility (in another set of risk ratios).

ChatGPT was used to assist with Excel table programming for odds ratios and chi squares; a member of the author team reviewed these suggestions before and after implementation to confirm their accuracy.

## Results

Table 1 provides descriptive data about the incidence of hypermobility and having an ONS profile pre-COVID in the full sample and three outcome categories: respondents who reported no Long COVID, respondents who reported Long COVID without high PESE, and respondents who reported Long COVID with high PESE. The percentages reflect the number of respondents in each subgroup divided by the total number of respondents in the outcome category.

**Table 1:**
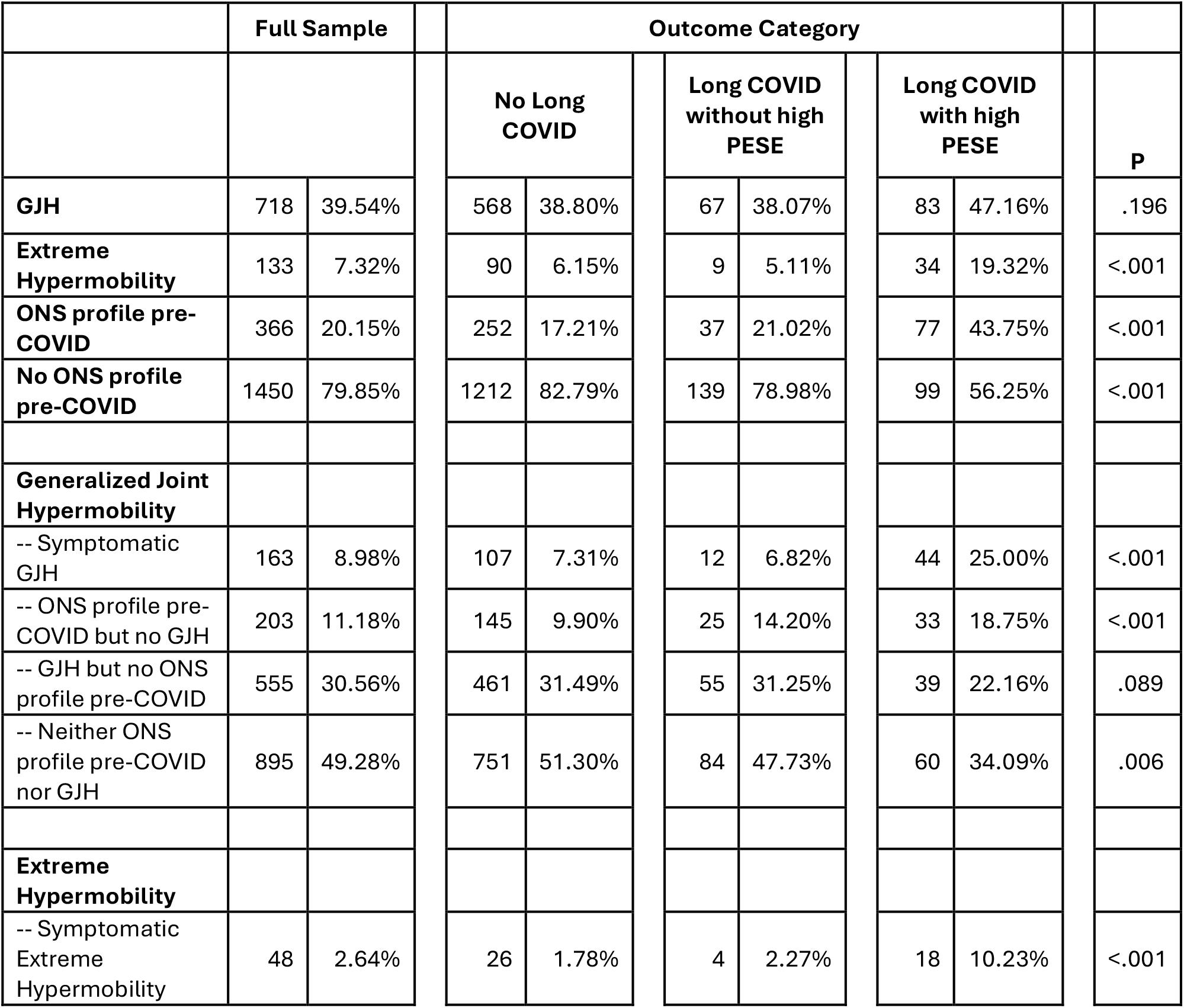
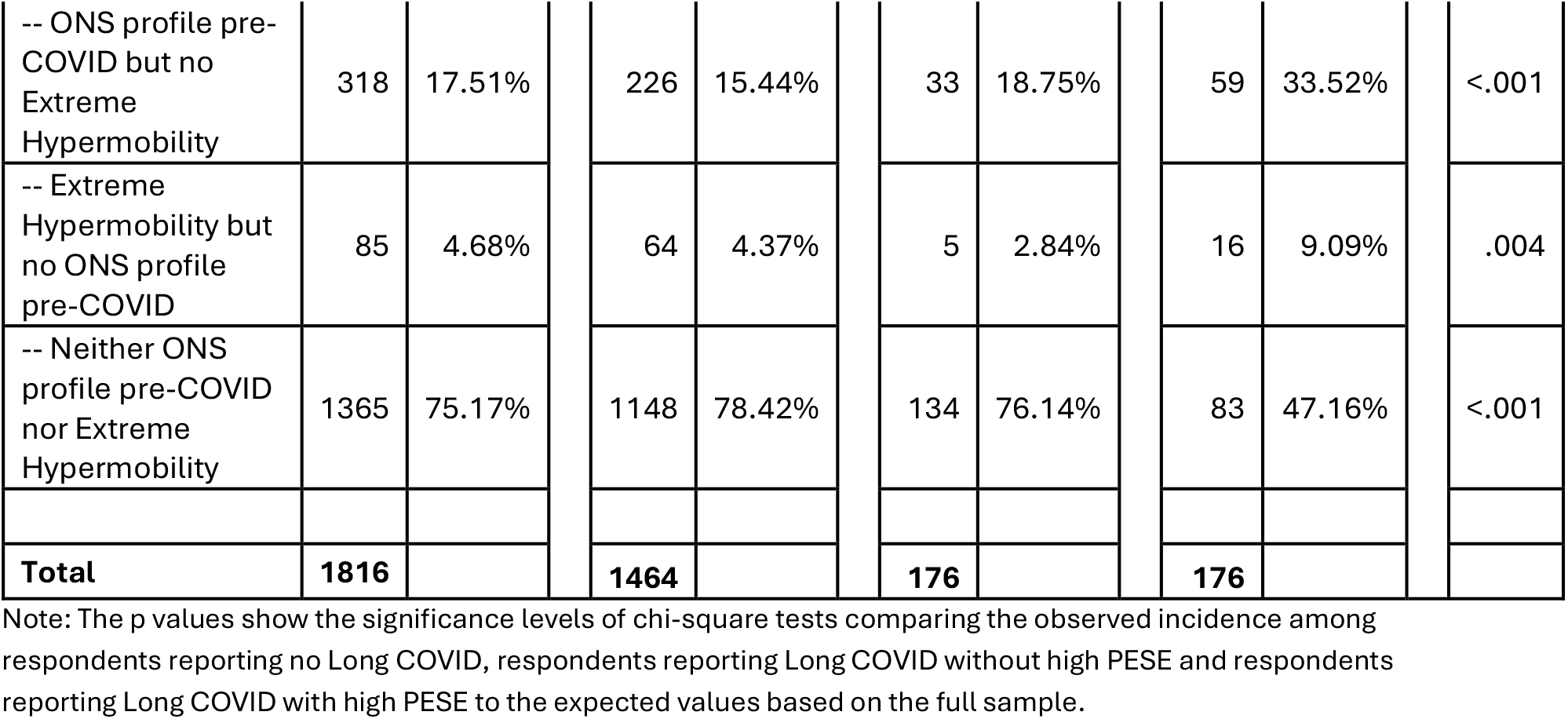
Incidence of GJH, symptomatic GJH, extreme hypermobility, and symptomatic extreme hypermobility.

As shown in Table 1, nearly 4 in 10 respondents reported GJH; the differences in incidence of GJH across the three outcome categories (columns) is not significant. However, with one other exception related to GJH, the differences across outcome categories (columns) for all other subgroups are significant. The last outcome category in particular stands out for showing comparatively high rates of both extreme hypermobility and an ONS profile pre-COVID among people reporting Long COVID with high PESE.

Table 1 also shows the interaction between hypermobility and having an ONS profile pre-COVID. Among people who did not report Long COVID, 7.31% have symptomatic GJH. This rises to 25.00% among those reporting Long COVID with high PESE. Among people who report no Long COVID, 1.78% had symptomatic extreme hypermobility; this rises to 10.23% among those reporting Long COVID with high PESE.

The rates of GJH, extreme hypermobility and an ONS profile pre-COVID among people with Long COVID without high PESE (third column) are not statistically different from the rates among people reporting no Long COVID (second column).

Table 2 reports the extent to which GJH, extreme hypermobility, an ONS profile pre-COVID, symptomatic GJH, and symptomatic extreme hypermobility are associated with respondents reporting Long COVID with high PESE. We show the results for the full sample and two subgroups: respondents reporting mild or no initial symptoms from (all) COVID-19 infection(s) and respondents reporting severe initial symptoms from at least one COVID-19 infection. Each regression in the table controls for age, sex at birth, number of infections (up to 3), and number of vaccine doses, factors previously shown to affect the risk of Long COVID.

**Table 2:**
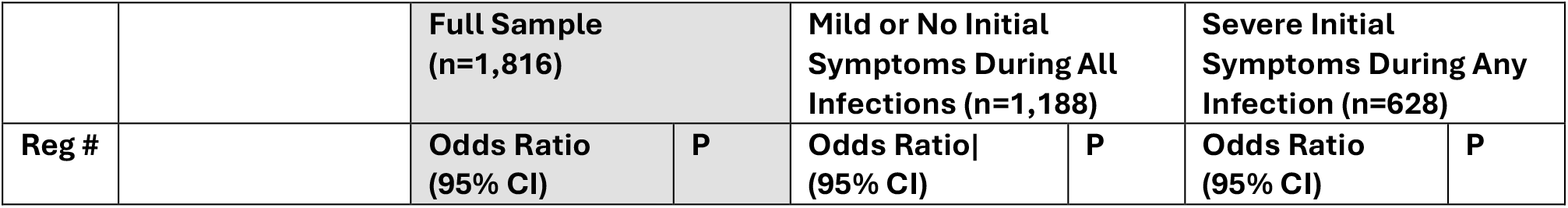
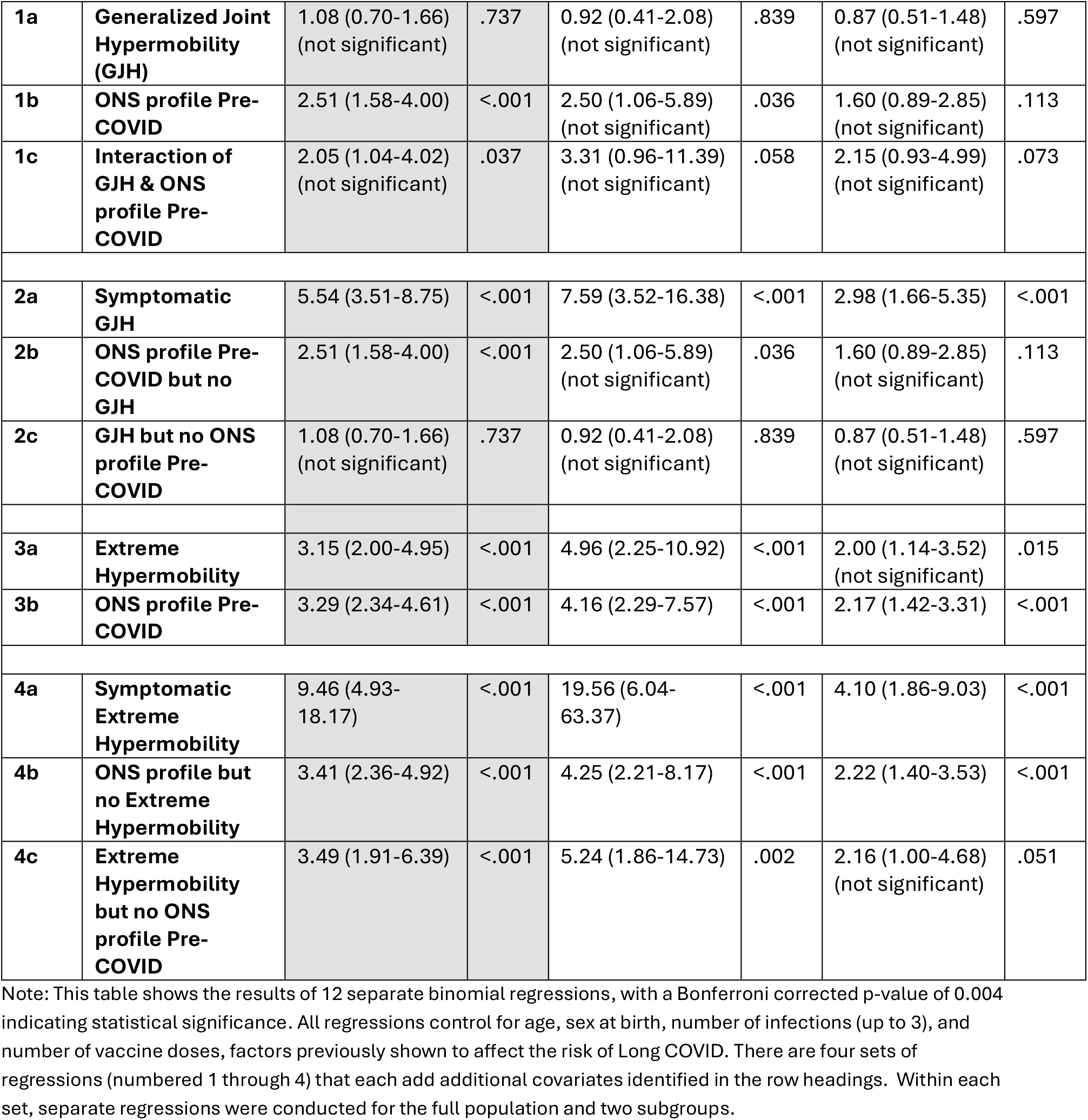
Binomial Logistic Regression Analysis Predicting Long COVID with high PESE.

|  |  | **Full Sample**<br>**(n=1,816)** |  | **Mild or No Initial**<br>**Symptoms During All**<br>**Infections (n=1,188)** |  | **Severe Initial**<br>**Symptoms During Any**<br>**Infection (n=628)** |  |
| --- | --- | --- | --- | --- | --- | --- | --- |
| **Reg #** |  | **Odds Ratio**<br>**(95% CI)** | **P** | **Odds Ratio**<br>**(95% CI)** | **P** | **Odds Ratio**<br>**(95% CI)** | **P** |
| **1a** | **Generalized Joint Hypermobility (GJH)** | 1.08 (0.70-1.66)<br>(not significant) | .737 | 0.92 (0.41-2.08)<br>(not significant) | .839 | 0.87 (0.51-1.48)<br>(not significant) | .597 |
| **1b** | **ONS profile Pre-COVID** | 2.51 (1.58-4.00) | <.001 | 2.50 (1.06-5.89)<br>(not significant) | .036 | 1.60 (0.89-2.85)<br>(not significant) | .113 |
| **1c** | **Interaction of GJH & ONS profile Pre-COVID** | 2.05 (1.04-4.02)<br>(not significant) | .037 | 3.31 (0.96-11.39)<br>(not significant) | .058 | 2.15 (0.93-4.99)<br>(not significant) | .073 |
| **2a** | **Symptomatic GJH** | 5.54 (3.51-8.75) | <.001 | 7.59 (3.52-16.38) | <.001 | 2.98 (1.66-5.35) | <.001 |
| **2b** | **ONS profile Pre-COVID but no GJH** | 2.51 (1.58-4.00) | <.001 | 2.50 (1.06-5.89)<br>(not significant) | .036 | 1.60 (0.89-2.85)<br>(not significant) | .113 |
| **2c** | **GJH but no ONS profile Pre-COVID** | 1.08 (0.70-1.66)<br>(not significant) | .737 | 0.92 (0.41-2.08)<br>(not significant) | .839 | 0.87 (0.51-1.48)<br>(not significant) | .597 |
| **3a** | **Extreme Hypermobility** | 3.15 (2.00-4.95) | <.001 | 4.96 (2.25-10.92) | <.001 | 2.00 (1.14-3.52)<br>(not significant) | .015 |
| **3b** | **ONS profile Pre-COVID** | 3.29 (2.34-4.61) | <.001 | 4.16 (2.29-7.57) | <.001 | 2.17 (1.42-3.31) | <.001 |
| **4a** | **Symptomatic Extreme Hypermobility** | 9.46 (4.93-18.17) | <.001 | 19.56 (6.04-63.37) | <.001 | 4.10 (1.86-9.03) | <.001 |
| **4b** | **ONS profile but no Extreme Hypermobility** | 3.41 (2.36-4.92) | <.001 | 4.25 (2.21-8.17) | <.001 | 2.22 (1.40-3.53) | <.001 |
| **4c** | **Extreme Hypermobility but no ONS profile Pre-COVID** | 3.49 (1.91-6.39) | <.001 | 5.24 (1.86-14.73) | .002 | 2.16 (1.00-4.68)<br>(not significant) | .051 |

Note: This table shows the results of 12 separate binomial regressions, with a Bonferroni corrected p-value of 0.004 indicating statistical significance. All regressions control for age, sex at birth, number of infections (up to 3), and number of vaccine doses, factors previously shown to affect the risk of Long COVID. There are four sets of regressions (numbered 1 through 4) that each add additional covariates identified in the row headings. Within each set, separate regressions were conducted for the full population and two subgroups.

For the full sample, the results show that having an ONS profile pre-COVID (OR 2.51, 95% CI 1.58 to 4.00) is associated with a significantly increased risk of Long-COVID with high PESE, but having GJH alone is not. The interaction between GJH and an ONS profile pre-COVID (OR 2.05, 95% CI 1.04 to 4.02) falls below the threshold for significance under the Bonferroni correction.

While the interaction term of GJH and an ONS profile pre COVID is not significant post-correction, it is nevertheless positive and reflected in the estimations of the risk of Long COVID with high PESE associated with symptomatic GJH in the second set of regressions, for which the OR is 5.54 (95% CI 3.51 to 8.75) for the full sample, 7.59 (95% CI 3.52 to 16.38) for respondents with mild or no initial symptoms from (all) COVID-19 infection(s) and 2.98 (95% CI 1.66 to 5.35) for respondents with severe initial symptoms from at least one COVID-19 infection.

Both extreme hypermobility and an ONS profile pre-COVID are robustly associated with Long COVID with high PESE in the third set of regressions, which cumulate to a very large, combined risk in the fourth set of regressions. Symptomatic extreme hypermobility is strongly associated with Long COVID with high PESE in both the full sample (OR 9.46, 95% CI 4.93 to 18.17) and among those with mild or no initial symptoms during (all) COVD-19 infection(s) (OR 19.56, 95% CI 6.04 to 63.37). Symptomatic extreme hypermobility is also associated with Long COVID with high PESE in the subgroup of respondents with severe initial symptoms from at least one COVID-19 infection(s) (OR 4.10, 95% CI 1.86 to 9.03), if less dramatically so.

Supplemental Table 1 shows the results of the same set of regressions using Long COVID (more generally) as the dependent outcome, rather than Long COVID with high PESE. The results are similar, though with lower ORs and the lack of significance (even before Bonferroni correction) of the interaction terms and for the subgroup with severe initial symptoms from at least one Long COVID infection(s).

Table 3 converts odds ratios to risk ratios to clarify the extent to which having symptomatic hypermobility increases the risk of Long COVID with high PESE. Both symptomatic GJH and symptomatic extreme hypermobility are associated with higher risks in the full sample and for both acute-infection-severity subgroups. In the full sample, having symptomatic GJH quadruples the risk of Long COVID with high PESE (RR 4.25, 95% CI 3.00 to 5.76) and having symptomatic extreme hypermobility increases the risk six-fold (RR 6.25, 95% CI 3.98 to 8.89). Among respondents with mild or no initial symptoms during (all) COVID-19 infection(s), having symptomatic GJH increases the risk six-fold (RR 6.33, 95% CI 3.27 to 11.17) and having symptomatic extreme hypermobility increases the risk 13-fold (RR 13.37, 95% CI 5.36 to 24.78). Among respondents with severe initial symptoms from at least one COVID-19 infection, both symptomatic GJH (RR 2.23, 95% CI 1.49 to 3.07) and symptomatic extreme hypermobility (RR 2.68, 95% CI 1.62 to 3.82) more than double the risk.

**Table 3:** Risk Ratios of Long COVID with High PESE associated with Symptomatic GJH and Symptomatic Extreme Hypermobility.

|  | Full Sample |  | Mild or No Initial Symptoms During All Infections |  | Severe Initial Symptoms During Any Infection |  |
| --- | --- | --- | --- | --- | --- | --- |
|  | Est. Incidence of Long COVID with High PESE | Risk Ratio | Est. Incidence of Long COVID with High PESE | Risk Ratio | Est. Incidence of Long COVID with High PESE | Risk Ratio |
| **Symptomatic GJH** | 28.5%<br>(20.1%-38.6%) | 4.25<br>(3.00-5.76) | 19.2%<br>(9.9%-33.9%) | 6.33<br>(3.27-11.17) | 37.9%<br>(25.4%-52.3%) | 2.23<br>(1.49-3.07) |
| **Neither ONS Profile Pre-COVID nor GJH** | 6.7% |  | 3.0% |  | 17.0% |  |
| **Symptomatic Extreme Hypermobility** | 38.0%<br>(24.2%-54.1%) | 6.25<br>(3.98-8.89) | 33.4%<br>(13.4%-61.9%) | 13.37<br>(5.36-24.78) | 39.2%<br>(23.7%-55.7%) | 2.68<br>(1.62-3.82) |
| Neither ONS<br>Profile Pre-COVID nor<br>Extreme<br>Hypermobility | 6.1% |  | 2.5% |  | 14.6% |  |

Note: the incidences of Long COVID with High PESE in the rows labelled “Neither ONS Profile Pre-COVID nor GJH” and “Neither ONS Profile Pre-COVID nor Extreme Hypermobility” are the unadjusted observed incidences in the data. The incidences of Long COVID with High PESE in the rows labelled “Symptomatic GJH” and “Symptomatic Extreme Hypermobility” are estimated based on the odds ratios reported in Table 2.

## Discussion

In this retrospective survey study, symptomatic hypermobility emerged as an unambiguous and clinically crucial predictor of Long COVID with post-exertional symptom exacerbation, a phenotype closely overlapping with ME. Extreme hypermobility and an ONS profile pre-COVID were each independently associated with increased risk, with a marked cumulative effect. In contrast, generalized joint hypermobility alone was not predictive of Long COVID with post-exertional symptom exacerbation, underscoring that it is the combination of hypermobility with multisystem symptom burden that generates the greatest vulnerability.

Strikingly, these associations were especially strong among individuals with mild or no symptoms during the acute phase of COVID-19 infection. This underscores the importance of looking beyond initial disease severity in identifying predictors of long-term outcomes. In people with mild or no initial symptoms who had symptomatic hypermobility, risk increased up to 13-fold, confirming the important role that host vulnerability plays in influencing Long COVID risk. This has direct implications for triage and early management: patients who appear “low risk” at point of infection may, in fact, require closer follow-up and screening for symptomatic hypermobility.

The pathogenesis of Long COVID in people with no or mild initial symptoms may well differ from that of people with severe initial symptoms, with the two groups representing separate phenotypes. Progress toward effective pharmacological treatments for the various forms of Long COVID will likely require stratification of the patient population according to Long COVID phenotype.^31^

Mechanistically, these findings are consistent with models in which baseline dysautonomia, vascular laxity, immune dysregulation, and impaired tissue resilience amplify the downstream impact of viral infection (as summarized in our earlier publication).^1^ These pathways have been implicated in both hypermobility syndromes and ME,^27^ and offer a plausible shared substrate for post-exertional symptom exacerbation. While causal inference is not possible here, the convergence of epidemiology and pathophysiology strengthens the case for targeted investigation.

More broadly, these results underscore the importance of considering the interaction between infection and pre-existing biological vulnerability. Symptomatic hypermobility is one such vulnerability: common, under-recognised, and clinically identifiable.

Further research is needed to understand the biological mechanisms of viral-induced illness to promote more effective and targeted treatments tailored to the disease pathways shared by groups of individuals with common vulnerabilities.

### Strengths and Limitations

As discussed in our previous publication,^1^ strengths of our survey include recruitment from a representative sample, use of a validated questionnaire (the 5PQ) that enables comparisons with other datasets and the inclusion of additional questions about hypermobility in other body parts; limitations include the use of self-reported retrospective data and the lack of controls for co-morbid conditions (which will be addressed in a subsequent analysis), and socioeconomic factors.

In addition to these points, it is important to emphasize the constraints of sample size for subgroup analysis. By definition, based on our methodology, no more than 10% of the sample has extreme hypermobility and only about 20% have an ONS profile pre-COVID-19; as a result, only 48 respondents have symptomatic extreme hypermobility. While this group is more likely to have Long COVID than other respondents, there are nevertheless only 22 who have Long COVID and 18 who have Long COVID with high PESE. The relatively small numbers in these subgroups help explain why there are wide confidence intervals around our estimates. It is important to replicate this analysis with a larger dataset that can enable more precise estimates. Conducting similar analyses with other datasets that include observations before individuals developed COVID-19 will help overcome the limitations imposed by reliance on retrospective data.

Another limitation specific to this analysis is that our proxy for an ME-type presentation of Long COVID only focuses on PESE and not the many other factors that go into a diagnosis of ME. This is driven largely by sample size, which prevented further refinement of this definition to include other factors that might have further constrained who meets the definition, and by the absence of direct observations or medical records to supplement the data we gathered through a retrospective survey.

One final limitation relates to the term “symptomatic hypermobility.” Many and perhaps most people with symptomatic hypermobility have one of the four orthostatic or neurocognitive symptoms used here to identify people with an ONS profile pre-COVID. But the term “symptomatic hypermobility” is also used to describe people with more localized symptoms, such as frequent joint subluxations. Our analysis of symptomatic hypermobility is limited to people in our sample with hypermobility and an ONS profile pre-COVID.

## Conclusion

The adverse continuing consequences of viral illnesses after the infectious period, as exemplified by Long COVID, remain a significant public health issue and a considerable burden to the individual and society. Unfortunately, with general interest in COVID-19 waning, millions are living with its long-term effects with few targeted treatment programs. This research adds to the growing literature underlining biological mechanisms of Long COVID, which increasingly implicate variant connective tissue as a contributing factor. Crucially, this research suggests that an inflammatory trigger, such as COVID-19, leads to more adverse outcomes in those with both hypermobility and a pre-existing orthostatic/neurocognitive symptom burden. Recognition of this cumulative effect in vulnerable individuals demands more targeted approaches to intervention strategies tailored to patients’ preexisting vulnerabilities and an improved understanding of the often-overlooked condition of symptomatic hypermobility.

## Data Availability

All data produced in the present study are available upon reasonable request to the authors.

## Author Contributions

JL, RAT, RR, JAE and LQ authors conceived, planned, and designed this study. JL, JAE, and LQ obtained funding. RAT and LQ collected data. JL conducted data analyses. JL, RAT, JAE, and LQ interpreted data. JL and RAT drafted the full manuscript. JAE, LQ, and RR edited the manuscript. JL, RAT, RR, JAE and LQ authors approved the final version. JE is the guarantor and attests that all listed authors meet authorship criteria and that no others meeting the criteria have been omitted.

## Acknowledgments

We are grateful to the individuals with Long COVID who provided feedback on the initial questionnaire and shared the pilot questionnaire via social media.

## Data availability

Data will be made available upon publication (link will be provided).

## Transparency Statement

The lead authors affirm that the manuscript is an honest, accurate, and transparent account of the study being reported; that no important aspects of the study have been omitted; and that any discrepancies from the study as planned (and, if relevant, registered) have been explained.

