## Supplemental Table 1 for "Symptomatic hypermobility as a risk factor for Long COVID with high post-exertional symptom exacerbation: further analysis of data from a retrospective online survey of adults in the United States and United Kingdom"

**Supplementary Materials**

### Contents

Supplemental Table 1: Binomial Logistic Regressions: Dependent Variable = Long COVID

*Supplemental Table 1: Binomial Logistic Regressions: Dependent Variable = Long COVID*

| | | Full Sample | | Mild or No Initial Symptoms During All Infections | | Severe Initial Symptoms During Any Infection | |
|---|---|---|---|---|---|---|---|
| **Reg #** | | **Odds Ratio (95% CI)** | **P** | **Odds Ratio\| (95% CI)** | **P** | **Odds Ratio (95% CI)** | **P** |
| **1a** | **Generalized Joint Hypermobility** | 1.26 (0.98-1.62) (Not significant) | .076 | 1.49 (1.02-2.19) | .041 | 0.90 (0.64-1.29) (Not significant) | .576 |
| **1b** | **ONS Profile Pre-COVID** | 2.26 (1.73-2.96) | <.001 | 2.29 (1.50-3.50) | <.001 | 1.58 (1.09-2.28) | .015 |
| **2a** | **Symptomatic GJH** | 3.06 (2.08-4.52) | <.001 | 3.80 (2.08-6.97) | <.001 | 1.62 (0.95-2.75) (Not significant) | .074 |
| **2b** | **ONS Profile Pre-COVID but no GJH** | 2.00 (1.39-2.87) | <.001 | 1.95 (1.09-3.48) | .024 | 1.22 (0.74-2.02) (Not significant) | .429 |
| **2c** | **GJH but no ONS Profile Pre-COVID** | 1.16 (0.86-1.56) (Not significant) | .329 | 1.36 (0.88-2.12) (Not significant) | .169 | 0.75 (.49-1.15) (Not significant) | .193 |
| **3a** | **Extreme Hypermobility** | 1.87 (1.25-2.80) | .002 | 2.37 (1.25-4.52) | .008 | 1.24 (0.73-2.10) (Not significant) | .435 |
| **3b** | **ONS Profile Pre-COVID** | 2.18 (1.67-2.86) | <.001 | 2.25 (1.47-3.45) | <.001 | 1.55 (1.07-2.25) | .020 |
| **4a** | **Symptomatic Extreme Hypermobility** | 4.56 (2.47-8.38) | <.001 | 8.49 (3.00-23.99) | <.001 | 1.99 (0.93-4.26) (Not significant) | .075 |
| **4b** | **ONS Profile Pre-COVID but no Extreme Hypermobility** | 2.12 (1.59-2.84) | <.001 | 2.06 (1.30-3.25) | .002 | 1.53 (1.03-2.28) | .036 |
| **4c** | **Extreme Hypermobility but no ONS Profile Pre-COVID** | 1.71 (1.01-2.91) | .047 | 1.77 (0.76-4.11) (Not significant) | .186 | 1.18 (0.58-2.42) (Not significant) | .644 |

Note: This table shows the results of 12 separate binomial regressions. All regressions control for age, sex at birth, number of infections (up to 3), and number of vaccine doses, factors previously shown to affect the risk of Long COVID. There are four sets of regressions (numbered 1 through 4) that each add additional covariates identified in the row headings. Within each set, separate regressions were conducted for the full population and two subgroups.
